# Early Mandated Social Distancing is a Strong Predictor of Reduction in Highest Number of New COVID-19 cases per Day within Various Geographic Regions

**DOI:** 10.1101/2020.05.07.20094607

**Authors:** Adnan I. Qureshi, M. Fareed K. Suri, Haitao Chu, Habibullah Khan Suri, Ayehsa Khan Suri

## Abstract

Mandated social distancing has been globally applied to limit the spread of corona virus disease 2019 (COVID-19) from highly pathogenic severe acute respiratory syndrome (SARS)-associated coronavirus 2 (SARS-CoV-2). The benefit of this community-based intervention in limiting COVID-19 has not been proven nor quantified. We examined the effect of timing of mandated social distancing on the rate of COVID-19 in 119 geographic regions derived from 41 states within United States and 78 countries. We found that highest number of new COVID-19 cases per day per million persons was significantly associated with total number of COVID-19 cases per million persons on the day before mandated social distancing (β=0.66, p<0.0001). Our findings suggest that the initiation of mandated social distancing for each doubling in number of existing COVID-19 cases would result in eventual peak with 58% higher number of COVID-19 infections per day. Subgroup analysis on those regions where the highest number of new COVID-19 cases per day have peaked increased β to .85 (p<0.0001). We demonstrate that initiating mandated social distancing at a 10 times smaller number of COVID-19 cases will reduce the number of daily new COVID-19 cases at peak by 80% highlighting the importance of this community-based intervention.

## Introduction

Quarantine and isolation are standard procedures to avoid transmission of infectious disease from infected to non-infected persons and have been used in numerous epidemics.**^1^** Social distancing has been another method for reducing frequency and closeness of contact between people in order to decrease the risk of transmission of disease. Social distancing has been used against influenza and corona virus disease 2019 (COVID-19) pandemics (caused by severe acute respiratory syndrome (SARS)-associated coronavirus 2 (SARS-CoV-2). Social distancing can be voluntary at individual level or mandated at a community level by governing authorities. Mandated social distancing comprises of a combination of travel restrictions, closure of non-essential group meeting venues (restaurants, schools, shops) and steps to avoid close contact at essential meeting venues (hospitals, food supply, pharmacies). Mandated social distancing is also referred to as societal lockdown and may have variable effect on disease transmission depending upon mode of transmission and ability to identify and isolate persons infected by the disease.**^2^** Critical analysis of mandated social distancing in 17 cities in United States during the 1918 pandemic (caused by H1N1 influenza A virus) found cities in which mandated social distancing were implemented at an early phase of the epidemic had peak death rates of 50% lower than those cities that did not implement such steps.**^3^** Although, the results from 1918 pandemic, influenzae pandemics, and SARS have been used to justify mandated social distancing in various parts of the world, limited analysis of the effect of mandated social distancing on COVID-19 pandemic are available. The value of mandated social distancing requires a critical assessment for each pandemic because of adverse psychological and health consequences on individuals**^4,5^** and financial effects on society.**^6^** We examine the effect of timing of mandated social distancing on the rate of COVID-19 infection in 119 geographic regions derived from 41 states within United States and 78 other countries.

## Methods

Daily cumulative COVID-19 case volumes for individual regions (countries and individual states of U.S.) since Jan 22,2020 are publicly available.**^7,8^** Mandated social distancing start dates for different regions have been compiled and are also available.**^9^** For this analysis, we included regions for which both mandated social distancing start dates and daily cumulative case volumes for COVID-19 were available. Except for the United States, we used national mandated social distancing start dates and national COVID-19 case volumes. For France, Denmark, Netherlands and United Kingdom, overseas regions were not included in calculation of national case volumes.

New COVID-19 cases per day were calculated from cumulative daily case volumes. We used 2019 population estimates for states in United States and other countries to calculate daily new and cumulative total COVID-19 case volumes per million persons residing within the region.**^10,11^** For further analysis, data was smoothed using moving average to remove daily fluctuations in reported COVID-19 cases. Smoothed data was plotted over raw data for all geographical regions to ensure that it is representative of raw data (appendix A). We excluded China as the curve was visually different from other regions and above methodology could not be reliably applied to that curve.

**Appendix A. 1.**
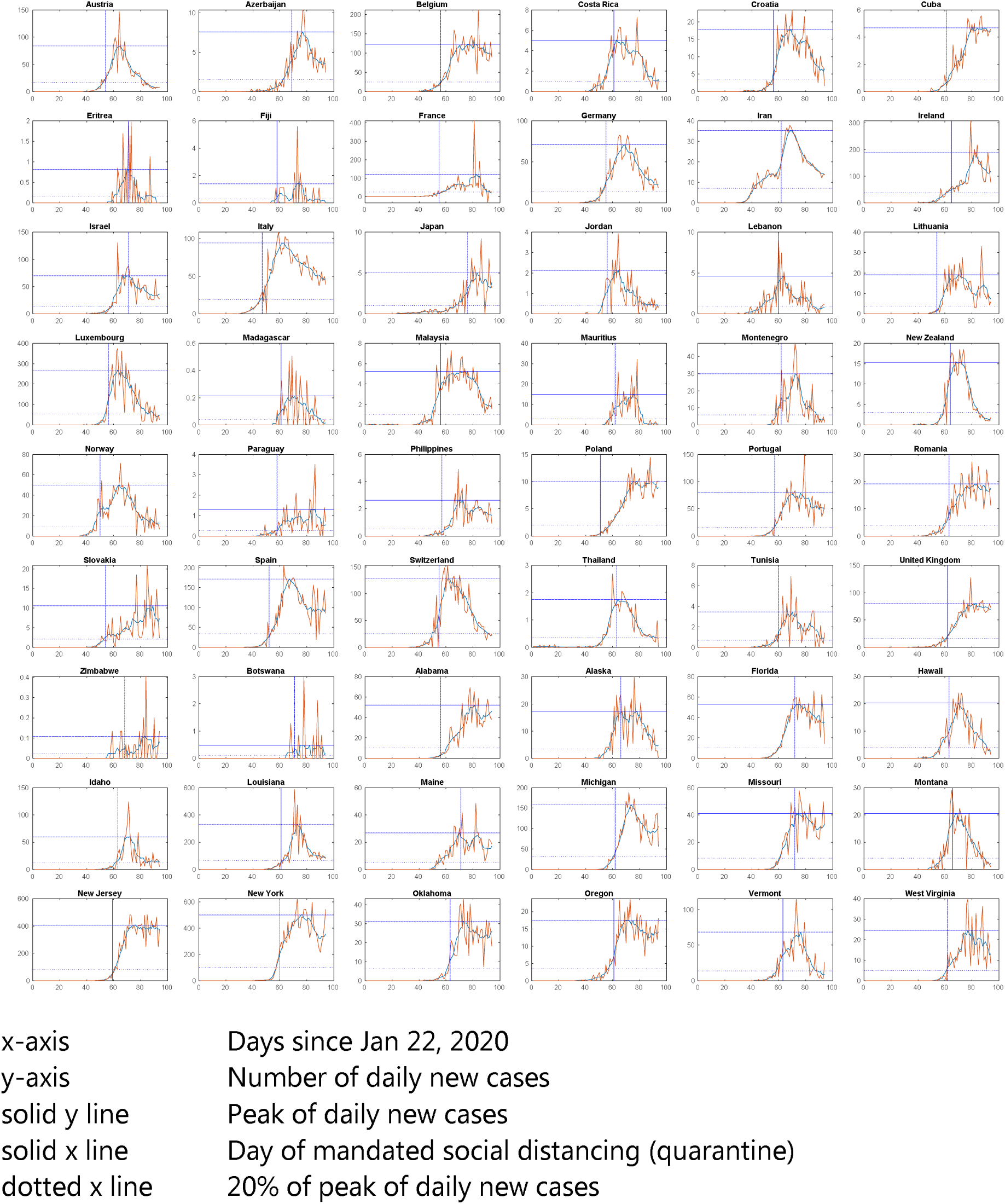
Geographical regions identified to have reached plateau in regard to number of new COVID-19 cases per day based on slope of last 13 days of new cases

**Appendix A. 2.**
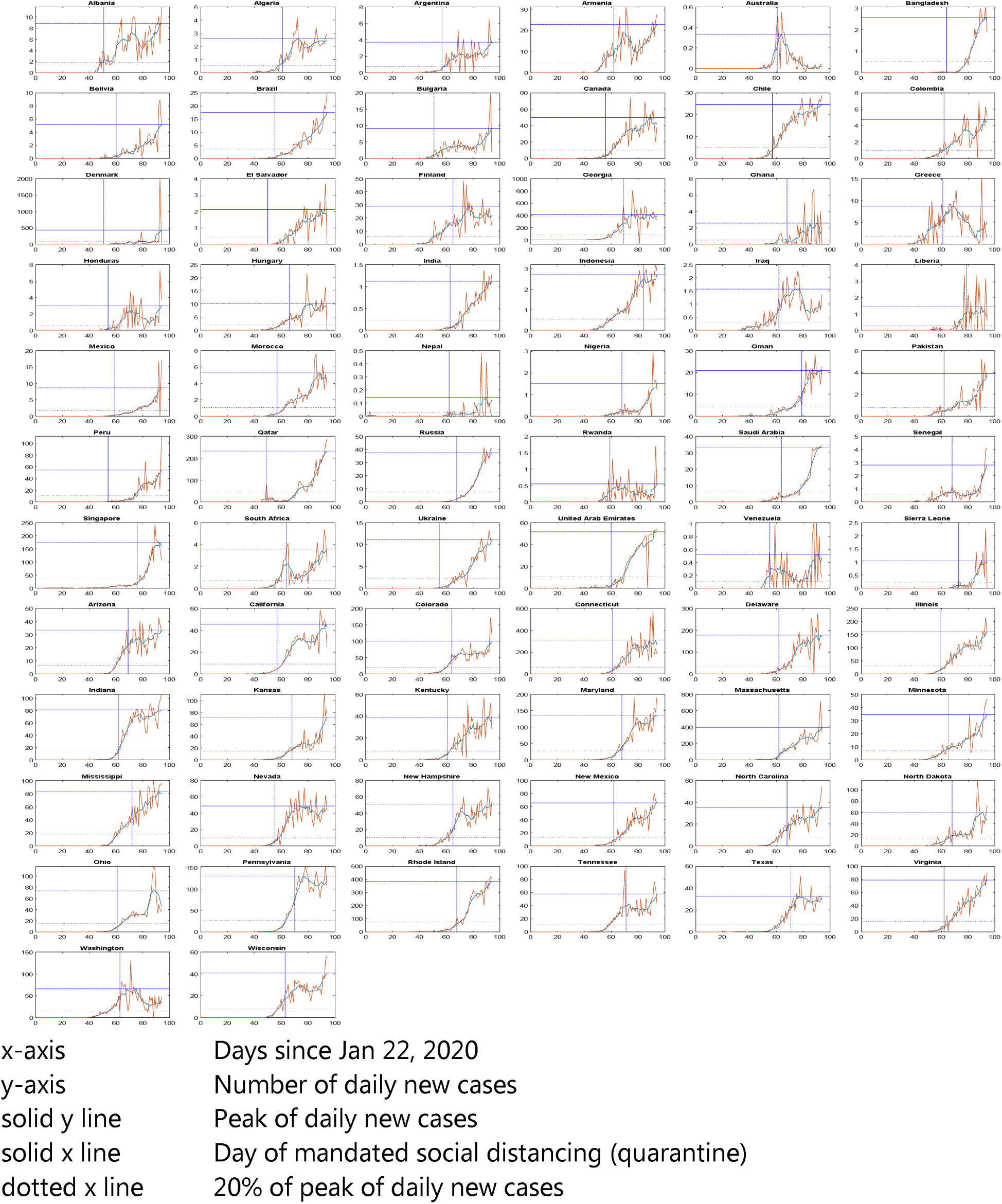
Geographical regions detected to be still trending up in regards to number of new COVID-19 cases per day based on slope of last 13 days of new cases

We used total number of COVID-19 cases per million on the day before mandated social distancing was implemented as the independent variable and predictor for our analysis. Peak of smoothed curve was used to determine highest number of new COVID-19 cases per day (expressed in per million persons) and used as the dependent variable. Due to the skewness in both the dependent and independent variables, log transformation was applied. To determine if the number of daily new cases have plateaued or are still increasing, we used linear regression for last 13 days. Last 13 days was selected after visually checking the trend for all geographic regions and repeating linear regression for various intervals ranging from 5 to 13 days. Linear positive trend for last 13 days (April 12^th^ to 25^th^) correlated best with visual interpretation of upward trend.

For all regression analysis we used log-transformed per million person values of highest number of new COVID-19 cases per day and total number of COVID-19 cases on the day before mandated social distancing. We used linear regression analysis to predict highest number of new COVID-19 cases per day using total number of COVID-19 cases on the day before mandated social distancing as predictor(Model A). Additional analysis of this association was performed after adjustment for the day mandated social distancing started in the course of COVID-19 pandemic -- calculated as number of days since 1/22/2020, log transformed population of geographic region and proportion of persons living in urban area (Model B).**^12,13^**

We repeated the analyses after classifying the geographic regions into those where the daily new COVID-19 case volume has plateaued and those regions where the COVID-19 was still increasing.

Using internet search, we manually abstracted individual elements of mandated social distancing for each of the geographical regions included in the analysis (Appendix B) and performed additional analysis after adjusting for these elements.

**Appendix B.**
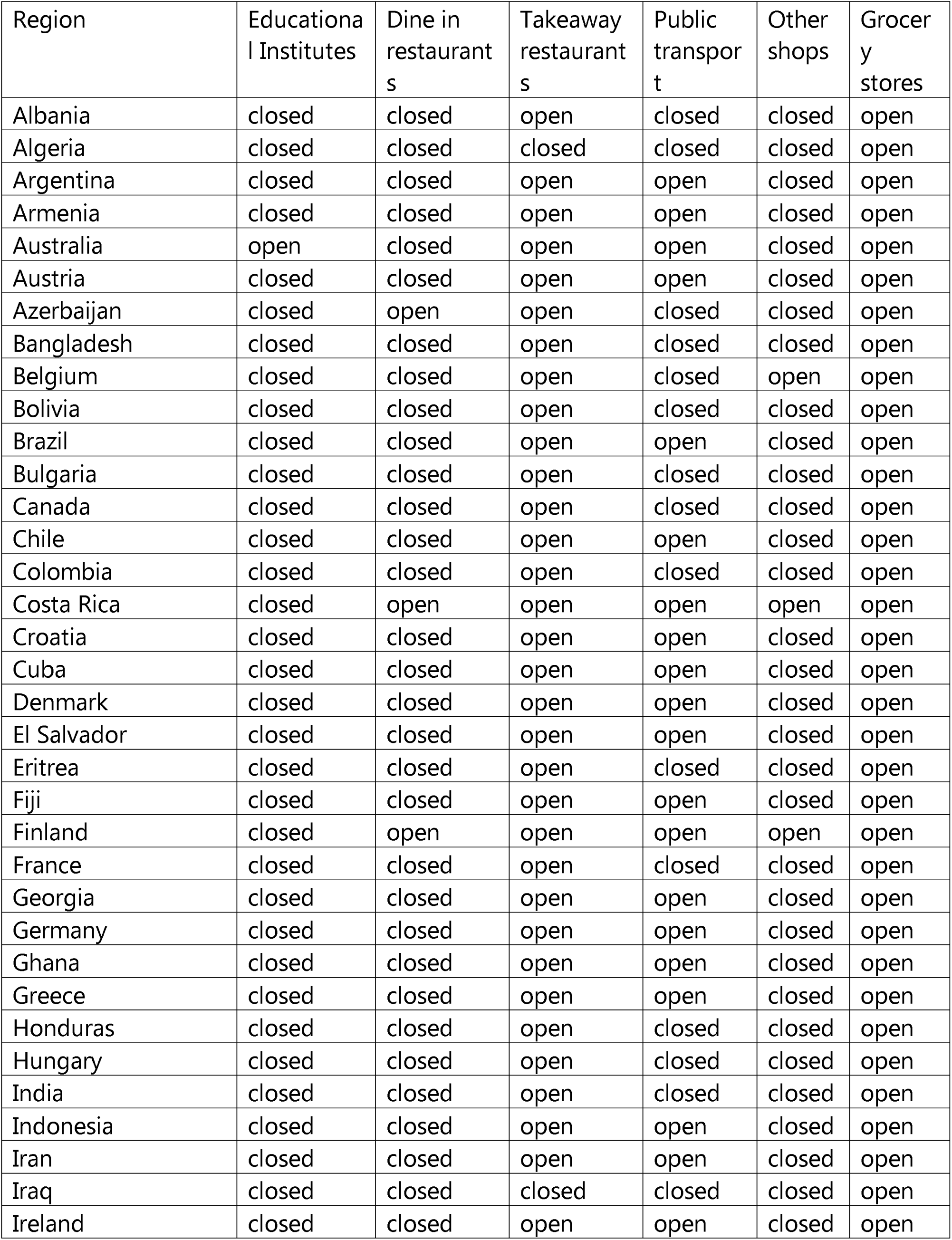
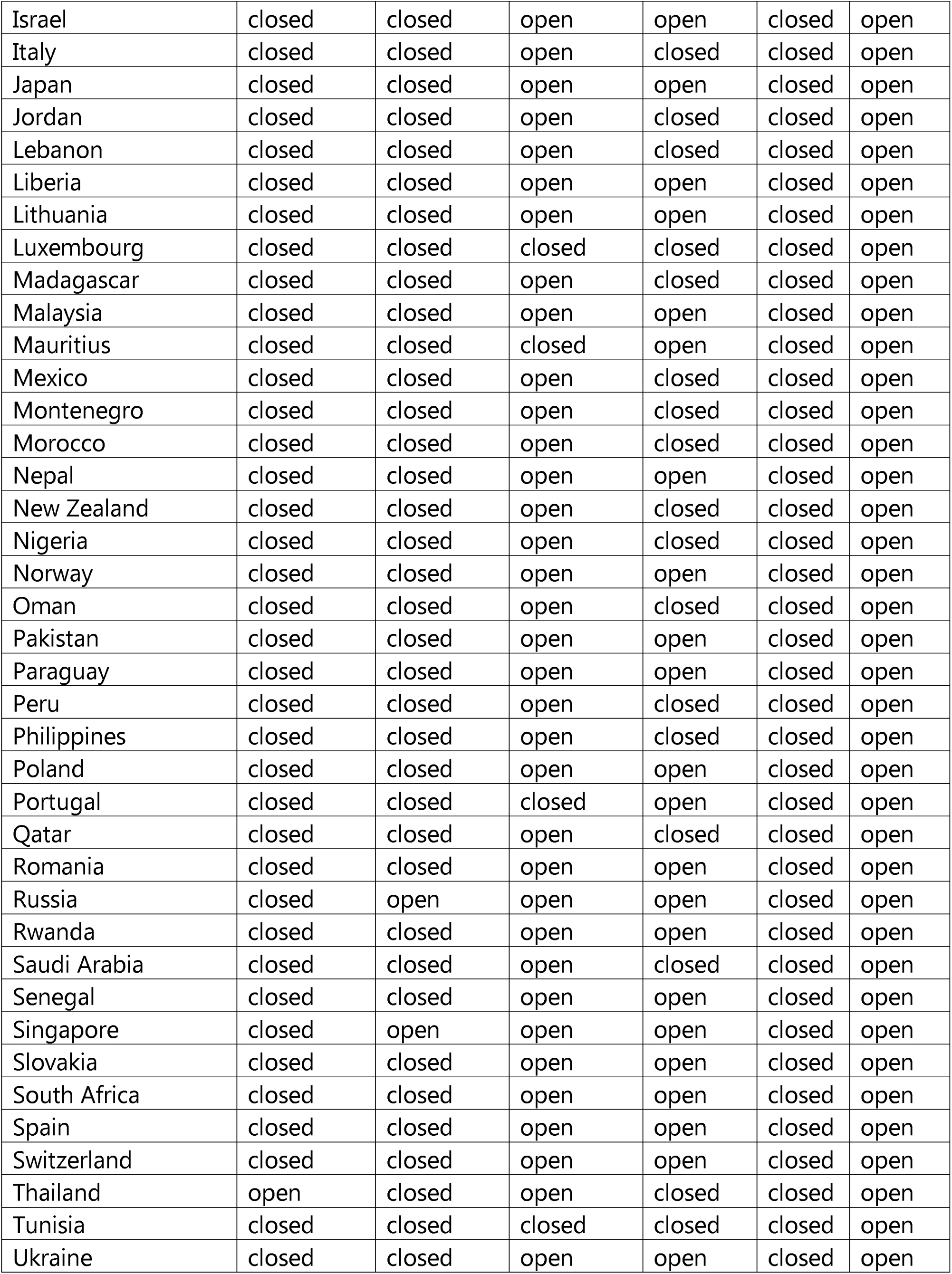
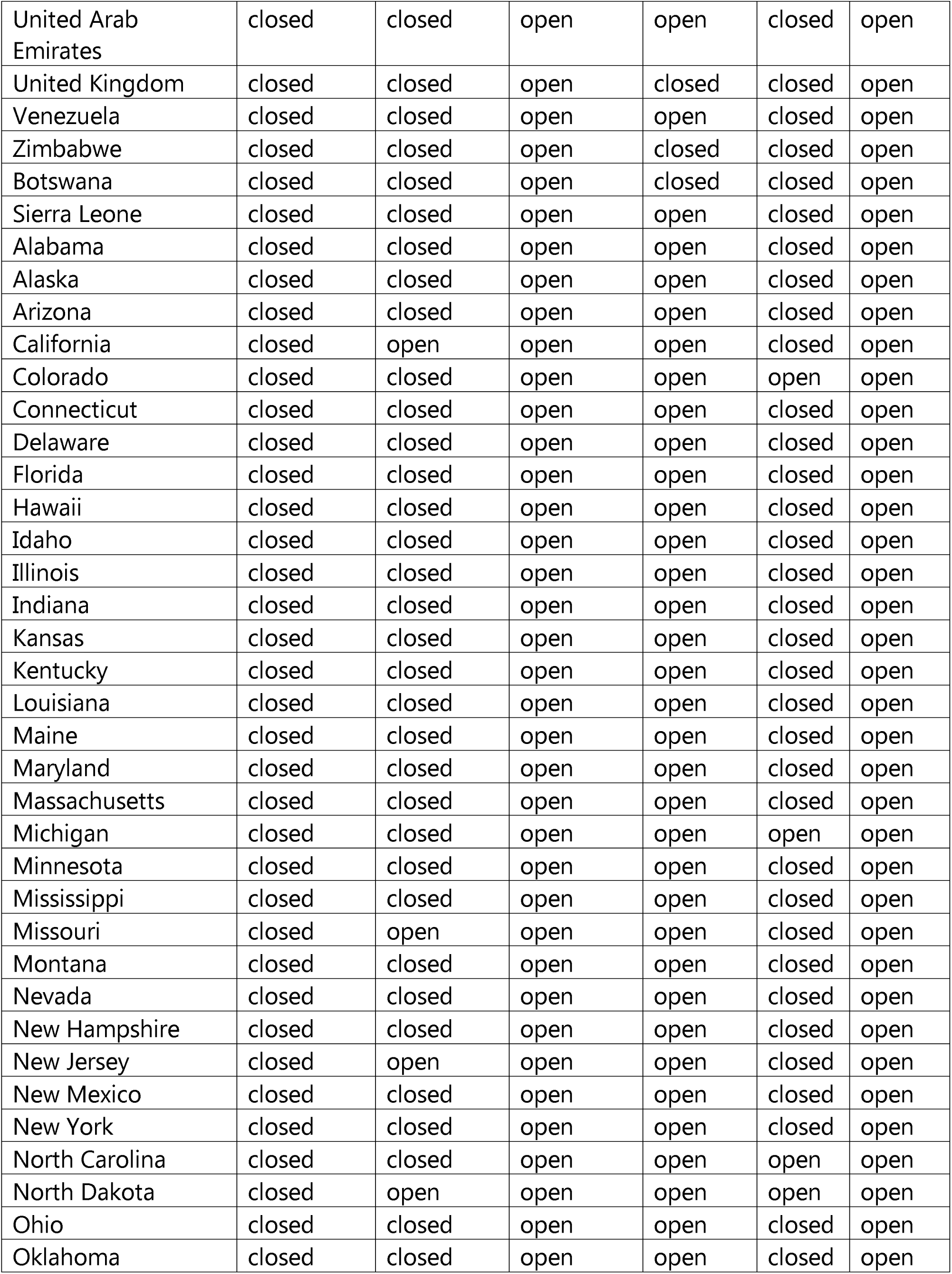
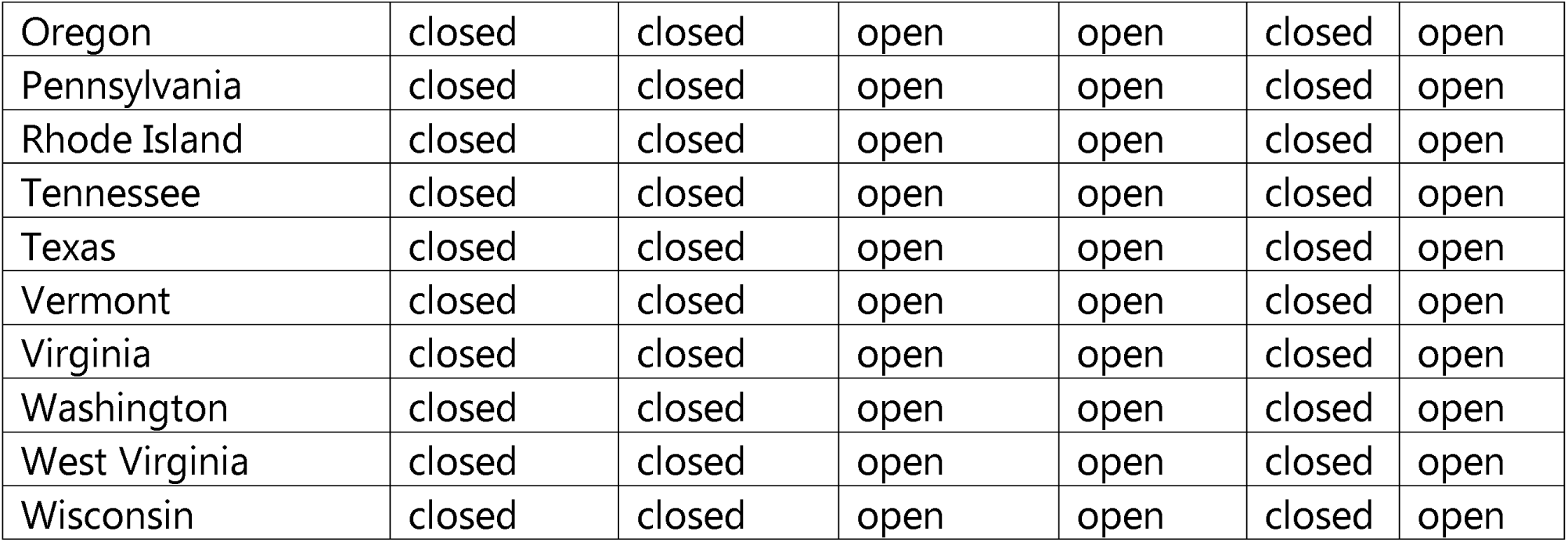
Elements of mandated social distancing in different regions

For regions where average (of last 5 days) daily new case volume has trended down to less than 20% of the peak daily new case volume (reached tail end), we performed linear regression analysis to predict the overall number of new COVID-19 cases per million from total number of COVID-19 cases per million persons on the day before mandated social distancing after log transformation of both variables.

## Results

Mandated social distancing dates of 85 countries and 42 states were available. Daily COVID-19 case volume data was available for 183 countries and all 52 states. After merger, both mandated social distancing starting dates and daily COVID-19 case census were available for 78 countries and 41 states. After excluding three regions for which the date of peak number of daily new cases was either after (Israel and Maine) or on the start day of mandated social distancing (Eritrea), number of days from the start date of mandated social distancing to the peak in daily new COVID-19 cases ranged from (1 – 45 days) (Figure 1).

**Figure 1:**
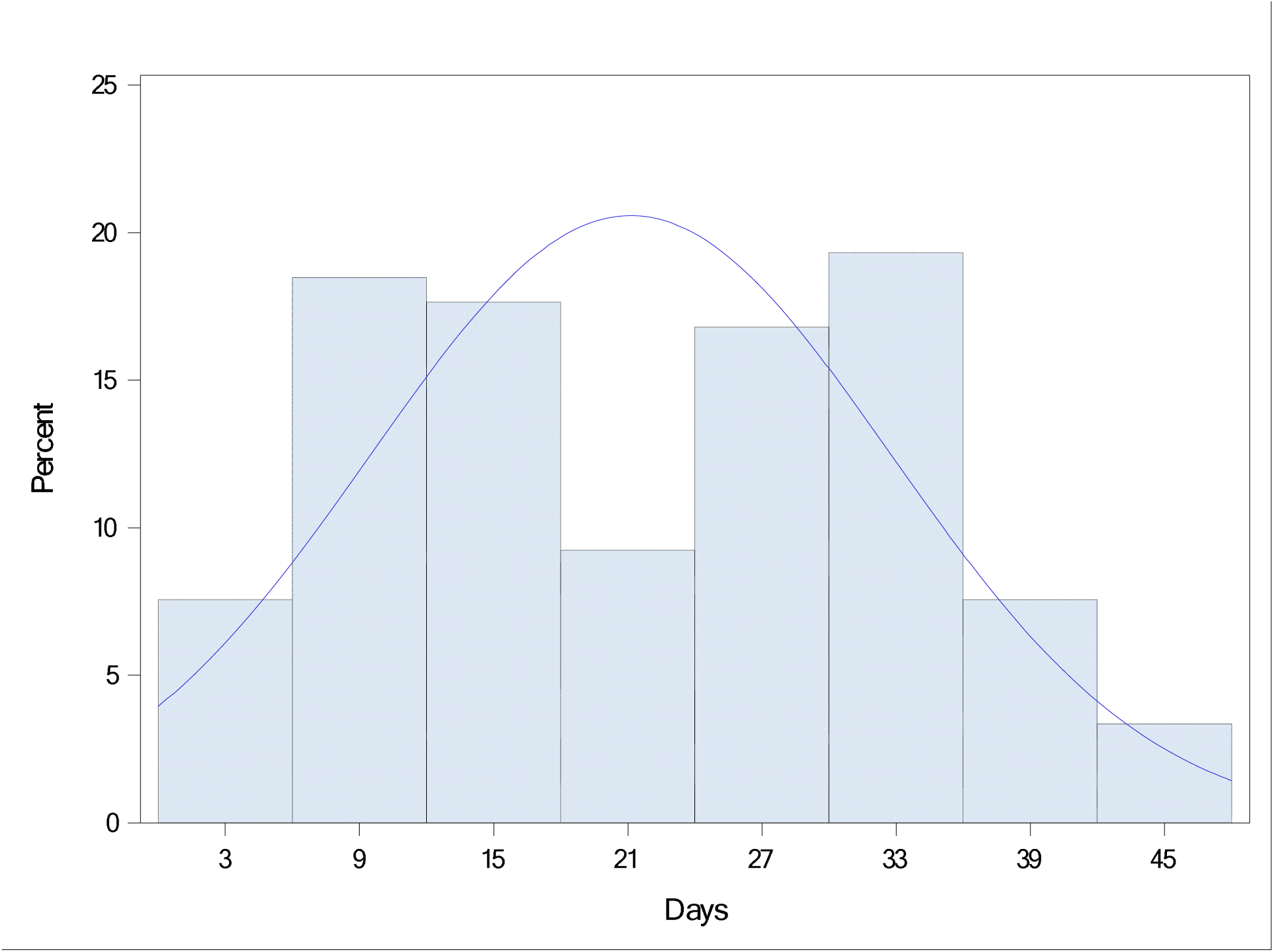
Interval (in days) between date of mandated social distancing and reaching the highest number of new COVID-19 cases per day

Mandated social distancing start dates within individual states of United States ranged from 3/17/2020 to 4/3/2020 and mandated social distancing dates of other countries ranged from 3/9/2020 to 4/15/2020. The total number of COVID-19 cases ranged from no cases to 1571 cases per million persons on the day before the start date of mandated social distancing, (Figure 2). Highest number of new COVID-19 cases per day ranged from 0.10 to 503 per million persons (Figure 3). There was clear trend towards association between total number of COVID-19 cases on the start date of mandated social distancing with highest number of new COVID-19 cases per day when plotted on logarithmic scale on scatter plot (Figure 5).

**Figure 3:**
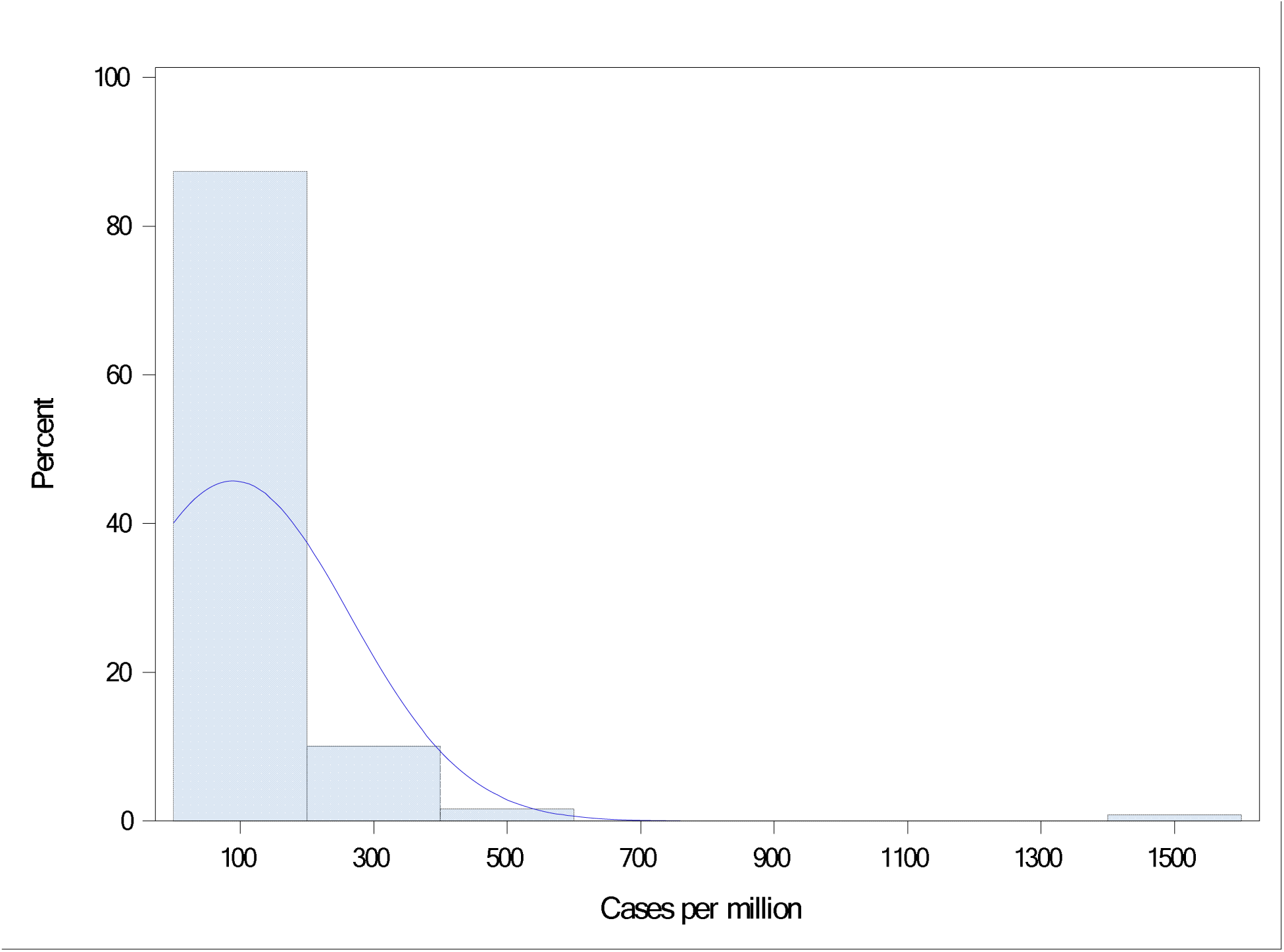
Distribution of total number of COVID-19 cases (per million population) on the day before start day of mandated social distancing

**Figure 4.**
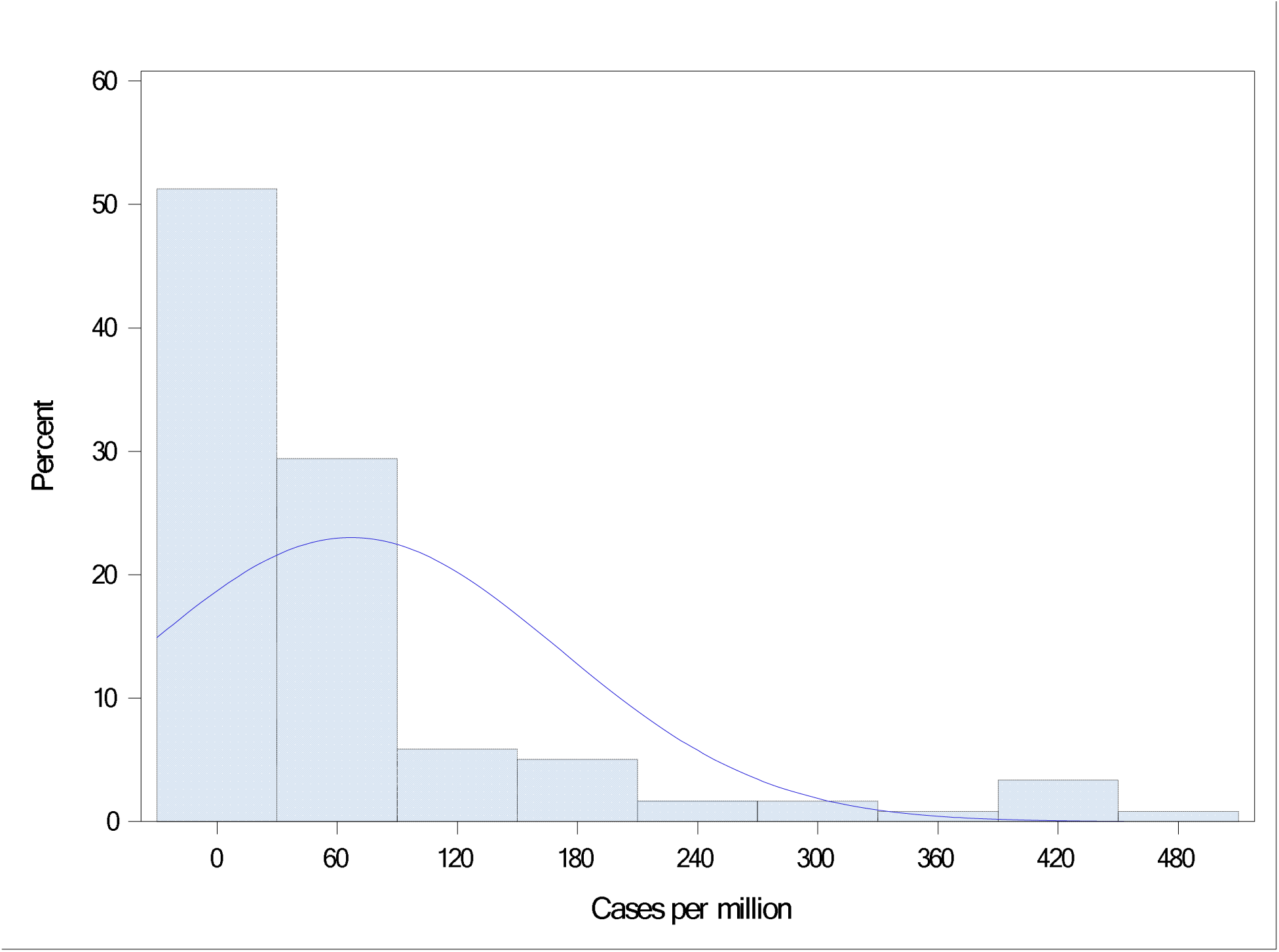
Distribution of highest number of new COVID-19 cases per day (per million population)

**Figure 5.**
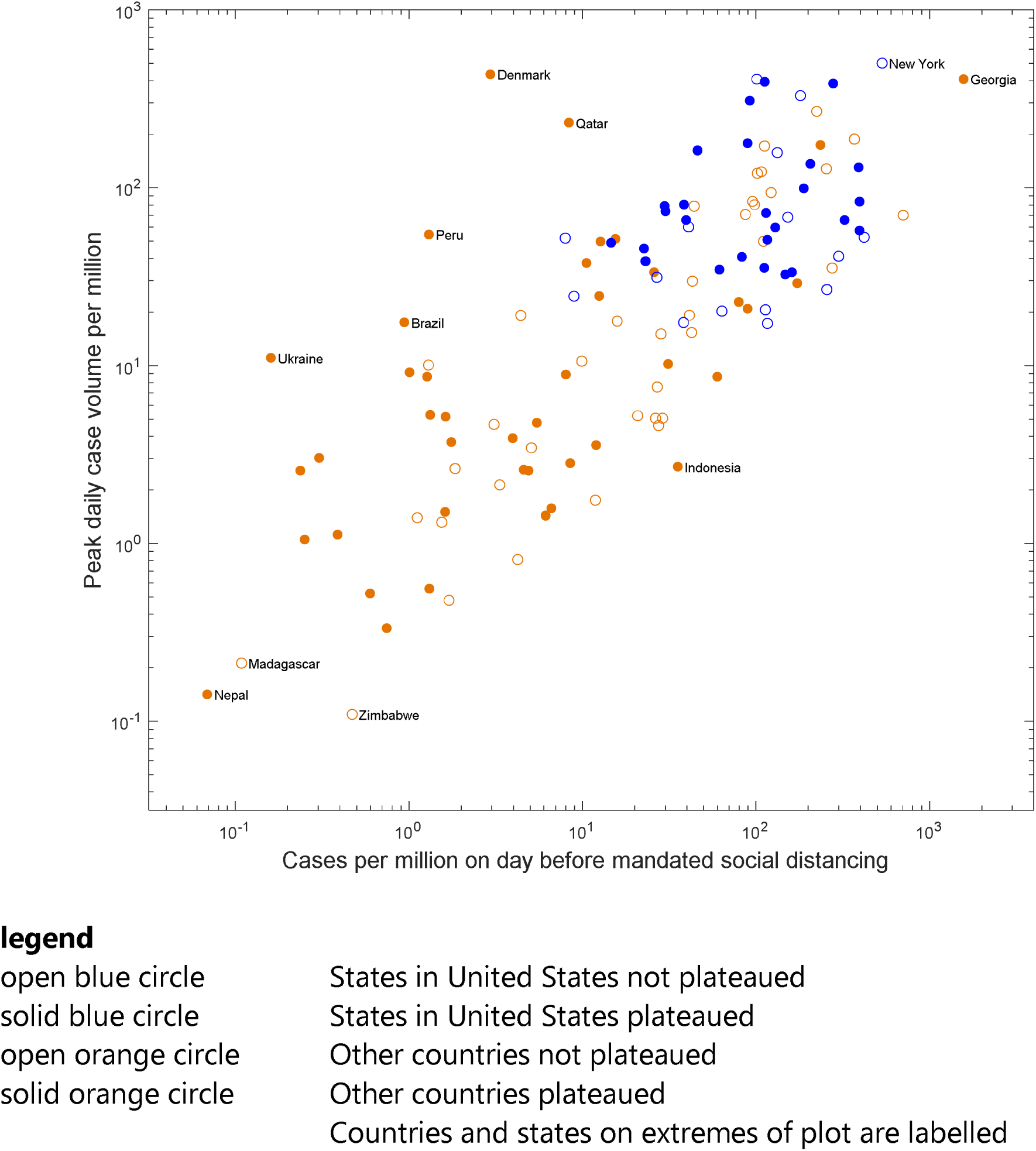
Relationship between total number of COVID-19 cases on day before start date of mandated social distancing and highest number of new COVID-19 cases per day on logarithmic scale

Results of linear regression analysis with different models are reported in Table 1. In Model A, highest number of new COVID-19 cases per day was significantly associated with total number of COVID-19 cases on the day before mandated social distancing (β=0.66, p<0.0001). Model B improved the adjusted R^2^ from 0.59 to 0.72 but did not change the β for total number of COVID-19 cases on the day before mandated social distancing. Subgroup analysis on those regions where the daily new COVID-19 cases have already peaked increased β for total number of COVID-19 cases on the day before mandated social distancing to .85 for both unadjusted and adjusted model (p<0.0001).

**Table 1:** Results of regression analysis predicting highest number of new COVID-19 cases per day

|  | All regions |  | Plateaued |  | Not-plateaued |  |
| --- | --- | --- | --- | --- | --- | --- |
| Total | 119 |  | 51 |  | 68 |  |
| States within United states | 41 |  | 15 |  | 26 |  |
| Other countries | 78 |  | 36 |  | 42 |  |
|  | Model A | Model B | Model A | Model B | Model A | Model B |
| F | 171.9 | 77.5 | 132.1 | 55.1 | 79.4 | 42.6 |
| Adjusted R2 | 0.59 | 0.72 | 0.72 | 0.81 | 0.54 | 0.71 |
| Constant | 10.1<br>(0.56) | 15.1<br>(1.65) | 11.8<br>(0.78) | 19.3<br>(2.15) | 9.6<br>(0.76) | 15.2<br>(2.56) |
| Log (cumulative case volume per million on the day before mandated social distancing) | 0.66***<br>(0.05) | 0.66***<br>(0.05) | 0.85***<br>(0.07) | 0.85***<br>(0.07) | 0.59***<br>(0.07) | 0.61***<br>(0.09) |
| Log (population of region) |  | -0.06<br>(0.07) |  | -0.08<br>(0.08) |  | -0.12<br>(0.11) |
| Day of mandated social distancing (from Jan 22,2020) |  | -0.09***<br>(0.02) |  | -0.1***<br>(0.02) |  | -0.08***<br>(0.02) |
| Percent of urban population in region |  | 0.02**<br>(0.006) |  | 0.001**<br>(0.008) |  | 0.02**<br>(0.009) |

standard errors are reported in parenthesis
\*, \*\*, \*\*\* indicates $p < 0.05$ , $p < 0.01$ , $p < 0.001$

Similar results from analysis for states within United States are reported in Table 1. There was less clear association between highest number of new COVID-19 cases per day and total number of COVID-19 cases on the day before mandated social distancing (β=0.3, p<0.001) in unadjusted model, but was stronger in adjusted model (β=0.72, p<0.0001). In a model (not shown in table) adjusted for only the day of mandated social distancing, the association between highest number of new COVID-19 cases per day and total number of COVID-19 cases on the day before mandated social distancing was strong (β=0.78, p<0.0001). Daily COVID-19 case volume had plateaued in only 13 states, and both unadjusted (Model A) and adjusted (Model B) association between highest number of new COVID-19 cases per day and total number of COVID-19 cases on the day before mandated social distancing was stronger in states where number of new cases had plateaued compared with states where the number of new COVID-19 case per day had not plateaued (Table 2).

**Table 2:** Results of regression analysis predicting highest number of new COVID-19 cases per day – United States

|  | All regions |  | Plateaued |  | Not-plateaued |  |
| --- | --- | --- | --- | --- | --- | --- |
| States in United States | 41 |  | 15 |  | 26 |  |
|  | Model A | Model B | Model A | Model B | Model A | Model B |
|  | 5.5 | 8.8 | 3.2 | 10.8 | 1.5 | 4.5 |
| Adjusted R2 | 0.1 | 0.44 | 0.13 | 0.74 | 0.02 | 0.36 |
| Constant | 7.1 (1.2) | 20 (4.01) | 7.9 (2.18) | 17.1 (5.15) | 6.1 (1.41) | 16.9 (5.85) |
| Log (cumulative case volume per million on the day before mandated social distancing) | 0.3*** (0.13) | 0.72*** (0.15) | 0.41*** (0.23) | 0.7*** (0.17) | 0.18*** (0.15) | 0.52*** (0.24) |
| Log (population of region) |  | 0.01 (0.12) |  | 0.5 (0.19) |  | -0.26 (0.13) |
| Day of mandated social distancing (from Jan 22,2020) |  | -0.15*** (0.04) |  | -0.2*** (0.04) |  | -0.08*** (0.06) |
| Percent of urban population in region |  | 0.009** (0.009) |  | -0.016** (0.013) |  | 0.023** (0.012) |

standard errors are reported in parenthesis
\*, \*\*, \*\*\* indicates $p < 0.05$ , $p < 0.01$ , $p < 0.001$

Internationally, there was a strong association between the highest number of new COVID-19 cases per day and total number of COVID-19 cases on the day before mandated social distancing both in the unadjusted or adjusted models (Table 3). This association was stronger for countries where the number of new COVID-19 cases per day has already plateaued (β=0.85, p<0.0001)

**Table 3:** Results of regression analysis predicting highest number of new COVID-19 cases per day - Other countries

|  | All regions |  | Plateaued |  | Not-plateaued |  |
| --- | --- | --- | --- | --- | --- | --- |
| Total | 78 |  | 36 |  | 42 |  |
|  | Model A | Model B | Model A | Model B | Model A | Model B |
|  | 87.6 | 42.9 | 129.5 | 58.3 | 23.5 | 14.3 |
| Adjusted R2 | 0.53 | 0.69 | 0.79 | 0.87 | 0.35 | 0.57 |
| Constant | 9.6 (0.8) | 12.8<br>(2.05) | 12.1<br>(0.86) | 18.4<br>(2.16) | 8.3<br>(1.32) | 12.3<br>(3.68) |
| Log (cumulative case volume per million on the day before mandated social distancing) | 0.63***<br>(0.07) | 0.6***<br>(0.06) | 0.88***<br>(0.08) | 0.83***<br>(0.07) | 0.51***<br>(0.1) | 0.55***<br>(0.12) |
| Log (population of region) |  | 0.02<br>(0.09) |  | -0.06<br>(0.08) |  | -0.02<br>(0.15) |
| Day of mandated social distancing (from Jan 22,2020) |  | -0.09***<br>(0.02) |  | -0.1***<br>(0.02) |  | -0.08***<br>(0.03) |
| Percent of urban population in region |  | 0.02**<br>(0.008) |  | 0**<br>(0.009) |  | 0.025**<br>(0.011) |

standard errors are reported in parenthesis
\*, \*\*, \*\*\* indicates $p < 0.05$ , $p < 0.01$ , $p < 0.001$

Addition of individual elements of mandated social distancing (closure of educational institutes, public transport, restaurants and other shops) did not affect the association between highest number of new COVID-19 cases per day and total number of COVID-19 cases on the day before mandated social distancing. Visually Australia was noted to have plateaued but based on a positive trend on last 13 days of regression it was classified as not plateaued. We repeated analysis of plateaued regions after manual addition of Australia and noticed no change in the above noted results.

For 17 regions (including 3 states of the United States), the daily new case volume was reduced to less than 20% of the peak daily new case volume. Log-transformed total number of cases was strongly predicted by total number of COVID-19 cases on the day before mandated social distancing (adjusted R-Square 0.87, F=112, (3=0.97, p<0.0001).

## Discussion

Our analysis confirmed the benefit and provided a quantitative estimate of value of mandated social distancing. Our findings suggest that the initiation of mandated social distancing for each doubling in number of existing COVID-19 cases would result in eventual peak with 60% higher number of COVID-19 infections per day. We found that that initiation of mandated social distancing at two folds higher number of existing COVID-19 cases would result in eventual peak with 58% higher number of COVID-19 cases (using (β of 0.66). If mandated social distancing is started when 100 persons are infected with COVID-19 and the highest number of cases is 1000 persons, initiating mandated social distancing when 200 persons are infected would increase the peak number of cases to 1580 persons. An example is mandated social distancing in New York which was initiated on day 61 when there were 10,356 cases. According to our analysis, if mandated social distancing was initiated on day 50 (142 cases), then the maximum number of cases per day would have been reduced by a factor of 16 (31 per million compared with 500 per million persons). We also identified what we consider is a spillover effect. There was blunting of effect in states within United States in the quantitative value of mandated social distancing when mandated social distancing was initiated later in the course of pandemic. We think that the blunting of effect was confounded by earlier mandated social distancing in surrounding states which resulted in mitigating the effect by reducing inflow of infected COVID-19 patients. This effect was not seen between countries where boundaries between countries may serve to insulate by restricting travel into the country. There are no restrictions between movement between states in United States enhancing the spillover effect.

Ferguson et al.**^1^** estimated that combining school and workplace closure with area quarantine and antiviral prophylaxis can result in 90% containment of infection (assuming the infection has R_0_ = 1.9), and when containment was initiated with less than 200 detected cases. The model was based on spread of H5N1 highly pathogenic avian influenza in wild and domestic poultry in Southeast Asia. Longini et al**^14^** modeled the Avian influenza A (subtype H5N1) outbreaks in Southeast Asia. The reported that local household quarantine is effective at containing the epidemic if R_0_ ≤ 2.1 but is not as effective at R_0_ = 2.4. However, a combination of 80% antiviral prophylaxis plus quarantine is effective at an R_0_ as high as 2.4, and adding previous vaccination makes antiviral prophylaxis plus quarantine even more effective. Both analyses mentioned that one of the reasons that limit the effect of mandated social distancing is the increase in contacts between household and neighborhood during social distancing which in highly infectious agents may offset the benefit. Ferguson et al.**^1^** assumed in their model that household and random contact rates increase by 100% and 50%, respectively, for individuals no longer able to attend school or work. Previous models have been based on H1N1 epidemiological experience. The R_0_ H1N1 flu has ranged between 1.25 in Canada**^15^**, 1.682 in China**^16^**, 1.96 in New Zealand**^17^**, 1.6 in Mexico**^18^** and 1.7 in United States**^19^**. One of the surprising findings is the benefit of mandated social distancing in COVID-19 pandemic despite the high infectivity of the SARS-CoV-2. The R_0_ of the SARS-CoV-2 infection was originally estimated between 2.2 and 2.7.**^20^**^-^**^25^** More recent data that suggest that the R_0_ of SARS-CoV-2 infection may be as high at 5.7.**^20^** The R_0_ of SARS-Co-V2 is higher than the threshold of 2.4 estimated by Longini et al**^14^** and 1.8 for new viral strains estimated by Ferguson et al.**^1^** where mandated social distancing will lose the beneficial effect.

There may be other reasons which may explain the beneficial effect of mandated social distancing in COVID-19 pandemic. Ridenhour et al.**^26^** had stressed upon the role of transmission rate, recovery rate, and size of population in the overall speed of the epidemic independent of Ro. Tang et al**^16^** had stressed upon the role of asymptomatic patients and those who are in prodromal period without symptoms in spread of pandemic H1N1 influenza in the province of Shaanxi. The beneficial effect of mandated social distancing may also be related to relatively long prodromal period and high proportion of asymptomatic SARS-CoV-2 infected patients. The time between transmission and symptoms ranges between 2–14 days for SARS-CoV-2.**^27^** Data on 468 COVID-19 transmission events reported in mainland China outside of Hubei Province found that in 59 (12.6%) of the 468 patients developed symptoms before the potential source developed symptoms suggesting that transmission occurred in the prodromal period.**^28^** There have been small case studies highlighting that the COVID-19 can be acquired from patients who are and will remain asymptomatic.**^29^**'**^31^** The estimated asymptomatic proportion of asymptomatic COVID-19 was 17.9% based on screening of travelers on board a cruise ship**^32^**, and 30.8% from data of Japanese citizens evacuated from Wuhan.**^33^** However, the viral loads in upper respiratory specimens appeared similar in symptomatic and asymptomatic persons.**^34^** We think that the beneficial effect of mandated social distancing may to related to reducing contact between asymptomatic person infected SARS-CoV-2. Another unique aspect of the SARS-CoV-2 is ability to persist on various surfaces and thus be transmitted by indirect contact from high touch surfaces.**^35,36^** SARS-CoV-2 can persist on plastic, stainless steel, copper and cardboard, and viable virus was detected up to 72 hours after application to these surfaces. The longest viability was on stainless steel and plastic; the estimated median half-life of SARS-CoV-2 was approximately 5.6 hours on stainless steel and 6.8 hours on plastic. Therefore, the mandated social distancing is likely to reduce contamination and transmission from high touch surfaces in society.

One of the limitations of our model is variability in policies pertaining to mandated social distancing and compliance to the policies in various geographic regions. The compliance to mandated social distancing introduced is an important factor in determining success of intervention.**^1^** There is also variability in exposure risk reduction among a given population as each individual within the population does not have the same chance of coming in contact with others.**^26^** There appears to be a difference according to age of the individuals**^37^** and population structure such as number of household, workplace, school, and community groups.**^38^** Differences in age and population structure between geographic regions may confound the results. There is also a confounding effect of case identification and isolation and robustness of testing for asymptomatic persons which may vary in various geographic units in our analysis. The Center for Disease Control and Prevention (CDC) concluded that the degree to which COVID-19 cases might go undetected or unreported varies in geographic regions because testing practices differ widely and might contribute significantly to the observed variations.**^39,40^**. For example, the state of New York (excluding NYC) reported administering 4.9 tests per 1,000 population, which was higher than the national average of 1.6 (CDC, unpublished data, March 25, 2020).

The variability in highest number of new cases per day that was not explained by our statistical models is likely due to variability in mandating social distancing in different regions. Although most of the businesses were closed during mandated social distancing, certain businesses like meat and poultry processing facilities were recognized as critical for infrastructure and permitted to continue work with precautions. Outbreaks in such places resulted in increasing number of new cases per day not explained by our model.**^41,42^** We also noted that in some regions (excluded from the analysis), highest number of new cases per day plateaued prior to mandated social distancing. This suggests that there may be other mechanisms that can reduce the number of new cases in certain regions. There were certain analyses which could not be performed for all the regions included in the analysis as the pandemic is ongoing. In subgroup analysis, it was clear that the relationship was strongest when the highest number of new cases per day had reached its peak. Some regions were still in the period where the highest number of new cases per day may continue to increase. The other issue was the total number of COVID-19 cases in a region which can only be determined after pandemic subsides. We did analyze17 regions where daily new cases have reached to less than 20% of the highest number of new cases per day observed (tail end of pandemic). There was a clear relationship between total cases before the start date of mandated social distancing and overall total number of case in the region indicating that early mandated social distancing also reduces the total number of COVID-19 infected persons affected overtime.

The value of mandated social distancing in reducing the spread of COVID-19 has been questioned at multiple levels due to widespread effects on individuals' wellbeing and sustenance and financial consequences on society. We demonstrate that initiating mandated social distancing when smaller number of COVID-19 cases are present will reduce the highest number of new cases per day and perhaps even the overall total number of COVID-19 cases in the region highlighting the importance of this community-based intervention.

## Data Availability

All data used for the analysis is either publicly available or included in the manuscript. I am in possession of all the data used in the manuscript and can provide upon request.

## References

1. Ferguson NM, Cummings DAT, Cauchemez S, et al. Strategies for containing an emerging influenza pandemic in Southeast Asia. Nature. 2005;437:209–214.

2. Ishola DA, Phin N. Could influenza transmission be reduced by restricting mass gatherings? Towards an evidence-based policy framework. J Epidemiol Glob Health. 2011;1:33–60.

3. Hatchett RJ, Mecher CE, Lipsitch M. Public health interventions and epidemic intensity during the 1918 influenza pandemic. Proc Natl Acad Sci U S A. 2007;104:7582–7587.

4. Shankar A, McMunn A, Demakakos P, Hamer M, Steptoe A. Social isolation and loneliness: Prospective associations with functional status in older adults. Health Psychol. 2017;36:179–187.

5. Shankar A, McMunn A, Banks J, Steptoe A. Loneliness, social isolation, and behavioral and biological health indicators in older adults. Health Psychol. 2011;30:377–385.

6. Correia S, Luck S and Verner El, Pandemics Depress the Economy, Public Health Interventions Do Not: Evidence from the 1918 Flu (March 30, 2020). Available at SSRN: https://ssrn.com/abstract=3561560 or http://dx.doi.org/10.2139/ssrn.3561560

7. Novel Coronavirus (COVID-19) Cases Data. https://data.humdata.org/dataset/novel-coronavirus-2019-ncov-cases. last accessed Apr 25, 2020

8. Coronavirus Source Data. https://ourworldindata.org/coronavirus-source-data. last accessed Apr 25, 2020

9. Global Covid-19 Lockdown Tracker. https://auravision.ai/covid19-lockdown-tracker/. last accessed Apr 20, 2020

10. State Population Totals and Components of Change: 2010-2019. https://www.census.gov/data/tables/time-series/demo/popest/2010s-state-total.html. last accessed Apr 20, 2020

11. Countries in the world by population. https://www.worldometers.info/world-population/population-by-country/. last accessed Apr 20, 2020

12. Census Urban and Rural Classification and Urban Area Criteria. https://www.census.gov/programs-surveys/geography/guidance/geo-areas/urban-rural/2010-urban-rural.html. last accessed Apr 20, 2020

13. Urban and Rural Populations. https://population.un.org/wup/Download/. last assessed Apr 22, 2020

14. Longini IM, Nizam A, Xu S, et al. Containing pandemic influenza at the source. Science. 2005;309:1083–1087.

15. Tuite AR, Greer AL, Whelan M, et al. Estimated epidemiologic parameters and morbidity associated with pandemic H1N1 influenza. CMAJ. 2010;182:131–136.

16. Tang S, Xiao Y, Yang Y, Zhou Y, Wu J, Ma Z. Community-based measures for mitigating the 2009 H1N1 pandemic in China. PLoS One. 2010;5:e10911.

17. Nishiura H, Wilson N, Baker MG. Estimating the reproduction number of the novel influenza A virus (H1N1) in a Southern Hemisphere setting: preliminary estimate in New Zealand. N Z Med J 2009;122:73–77.

18. Fraser C, Donnelly CA, Cauchemez S, et al. Pandemic potential of a strain of influenza A (H1N1): early findings. Science. 2009;324:1557–1561.

19. White LF, Wallinga J, Finelli L, et al. Estimation of the reproductive number and the serial interval in early phase of the 2009 influenza A/H1N1 pandemic in the USA. Influenza Other Respir Viruses. 2009;3:267–276.

20. Wu JT, Leung K, Leung GM. Nowcasting and forecasting the potential domestic and international spread of the 2019-nCoV outbreak originating in Wuhan, China: a modelling study. Lancet. 2020;395:689–697.

21. Li Q, Guan X, Wu P. Early Transmission Dynamics in Wuhan, China, of Novel Coronavirus-Infected Pneumonia. N Engl J Med. 2020;382:1199–1207.

22. Du Z, Wang L, Cauchemez S, et al. Risk for Transportation of Coronavirus Disease from Wuhan to Other Cities in China. Emerg Infect Dis. 2020;26:1049–1052.

23. Riou J, Althaus CL. Pattern of early human-to-human transmission of Wuhan 2019 novel coronavirus (2019-nCoV), December 2019 to January 2020. Euro Surveill. 2020;25: doi, forthcoming 2020.

24. Yuan J, Li M, Lv G, Lu ZK. Monitoring Transmissibility and Mortality of COVID-19 in Europe. Int J Infect Dis. 2020: doi, forthcoming 2020.

25. Sanche S, Lin YT, Xu C, Romero-Severson E, Hengartner N, Ke R. High Contagiousness and Rapid Spread of Severe Acute Respiratory Syndrome Coronavirus 2. Emerg Infect Dis. 2020;26: doi, forthcoming 2020.

26. Ridenhour B, Kowalik JM, Shay DK. Unraveling R0: considerations for public health applications. Am J Public Health. 2014;104:e32-e41.

27. Li C, Ji F, Wang L, et al. Asymptomatic and Human-to-Human Transmission of SARS-CoV-2 in a 2-Family Cluster, Xuzhou, China. Emerg Infect Dis. 2020;26: doi, forthcoming 2020.

28. Lauer SA, Grantz KH, Bi Q, et al. The Incubation Period of Coronavirus Disease 2019 (COVID-19) From Publicly Reported Confirmed Cases: Estimation and Application. Ann Intern Med. 2020: doi, forthcoming 2020.

29. Rothe C, Schunk M, Sothmann P, et al. Transmission of 2019-nCoV Infection from an Asymptomatic Contact in Germany. N Engl J Med. 2020;382:970–971.

30. Du Z, Xu X, Wu Y, Wang L, Cowling BJ, Meyers LA. Serial Interval of COVID-19 among Publicly Reported Confirmed Cases. Emerg Infect Dis. 2020;26: doi, forthcoming 2020.

31. Bai Y, Yao L, Wei T, et al. Presumed Asymptomatic Carrier Transmission of COVID-19. JAMA. 2020: doi, forthcoming 2020.

32. Chan JF-W, Yuan S, Kok K-H, et al. A familial cluster of pneumonia associated with the 2019 novel coronavirus indicating person-to-person transmission: a study of a family cluster. Lancet. 2020;395:514–523.

33. Nishiura H, Kobayashi T, Suzuki A, et al. Estimation of the asymptomatic ratio of novel coronavirus infections (COVID-19). Int J Infect Dis. 2020: doi, forthcoming 2020.

34. Zou L, Ruan F, Huang M, et al. SARS-CoV-2 Viral Load in Upper Respiratory Specimens of Infected Patients. N Engl J Med. 2020;382:1177–1179.

35. Santarpia J, Rivera D, Herrera V, Morwitzer M, Creager H, Santarpia G, et al. Transmission Potential of SARS-CoV-2 in Viral Shedding Observed at the University of Nebraska Medical Center. pre print. doi: https://doi.org/10.1101/2020.03.23.20039446

36. van Doremalen N, Bushmaker T, Morris DH, et al. Aerosol and Surface Stability of SARS-CoV-2 as Compared with SARS-CoV-1. N Engl J Med. 2020;382:1564–1567.

37. Nokes DJ, Anderson RM. The use of mathematical models in the epidemiological study of infectious diseases and in the design of mass immunization programmes. Epidemiol Infect. 1988;101:1–20.

38. Pellis L, Ferguson NM, Fraser C. Threshold parameters for a model of epidemic spread among households and workplaces. J R Soc Interface. 2009;6:979–987.

39. CDC COVID-19 Response Team. Geographic Differences in COVID-19 Cases, Deaths, and Incidence - United States, February 12-April 7, 2020. MMWR Morb Mortal Wkly Rep. 2020;69:465–471.

40. Verity R, Okell LC, Dorigatti I, et al. Estimates of the severity of coronavirus disease 2019: a model-based analysis. Lancet Infect Dis. 2020: doi, forthcoming 2020.

41. Meat and Poultry Processing Workers and EmployersInterim Guidance from CDC and the Occupational Safety and Health Administration (OSHA). https://www.cdc.gov/coronavirus/2019-ncov/community/organizations/meat-poultry-processing-workers-employers.html. last assessed May 3, 2020

42. COVID-19 Among Workers in Meat and Poultry Processing Facilities? 19 States, April 2020. https://www.cdc.gov/mmwr/volumes/69/wr/mm6918e3.htm?s_cid=mm6918e3_w#T1_down. last assessed May 3, 2020

